# Novelty-induced frontal-STN networks in Parkinson’s disease

**DOI:** 10.1101/2021.06.24.21259502

**Authors:** Rachel C Cole, Arturo I Espinoza, Arun Singh, Joel I Berger, James F Cavanagh, Jan R Wessel, Jeremy D Greenlee, Nandakumar S Narayanan

## Abstract

Novelty detection is a primitive subcomponent of cognitive control that is deficient in Parkinson’s disease (PD) patients with cognitive dysfunction. Here, we studied novelty-response mechanisms in PD. In participants with PD, we recorded from cortical circuits with scalp-based electroencephalography (EEG) and from subcortical circuits using intraoperative neurophysiology during surgeries for implantation of deep-brain stimulation (DBS) electrodes. We report three major results. First, novel auditory stimuli triggered midfrontal low-frequency rhythms; of these, 1-4 Hz “delta” rhythms were linked to novelty-associated slowing whereas 4-7 Hz “theta” rhythms were specifically attenuated in PD. Second, 32% of subthalamic nucleus (STN) neurons were response-modulated; nearly all (94%) of these were also modulated by novel stimuli. Third, response-modulated STN neurons were coherent with midfrontal 1-4 Hz activity. These findings link scalp-based measurements of neural activity with neuronal activity in the STN. Our results provide insight into midfrontal cognitive control mechanisms and how purported hyperdirect fronto-basal ganglia circuits evaluate new information.

## INTRODUCTION

New information requires careful consideration. Indeed, when the brain evaluates novel information, it recruits cognitive-control processes reflected by midfrontal low-frequency “delta” (1-4 Hz) and “theta” (4-7) Hz activity (Cavanagh & Frank, 2014; Solís-Vivanco et al., 2018). In the prefrontal cortex, low-frequency activity encodes cognitive functions (Cavanagh et al., 2012; Cooper et al., 2019; Messel et al., 2021), as well as engages single neurons in subcortical regions of the brain (Kim & Narayanan, 2019; Mikell et al., 2014; Narayanan et al., 2013). A better understanding of the systems-level interactions underlying control may contribute to better treatment of disorders associated with control deficits, such as Parkinson’s disease (PD). While PD is primarily associated with motor control issues, there are also profound disturbances in cognitive control. To elucidate the arc between endogenous elicitation and exogenous exertion, we examined the systems-level interactions between midfrontal field potentials and subcortical neurons in PD patients.

We utilized the well-known oddball task to evoke novelty-incuded control (Parmentier, 2014; Parmentier & Hebrero, 2013). In this task, participants must respond to a commonly-occurring, standard stimulus on most trials. Novel stimuli can trigger a cascade of processes including the orienting response (Sokolov, 1962), motor effects, and cognitive control (Fan, 2014). When novel distractors interrupt a primary task, responses are slower and less accurate (Parmentier & Hebrero, 2013; Wessel & Huber, 2019). When cognitive control is appropriately engaged, this novelty-induced distraction can be reduced (e.g. a control rule could determine that the distractors are irrelevant and thus should be ignored) (Parmentier & Hebrero, 2013). Thus, novelty-induced orienting signals a generic need for control.

In electroencephalography (EEG) studies, novel stimuli and the subsequent orienting response evoke characteristic event-related potential (ERP) signatures in the midfrontal cortex (Debener et al., 2005; Polich, 2007; Ranganath & Rainer, 2003). Cortical responses to novel stimuli involve midfrontal low-frequency oscillations in the delta (1-4 Hz) and the theta (4-7 Hz) range (Cavanagh et al., 2012; Wessel et al., 2016). Cavanagh and Frank (2014) proposed that these midfrontal oscillations reflect the recruitment of cognitive-control processes, however, the precise bands and topography of novelty-related slowing, particularly in PD patients, is undetermined. Frontal activity may serve as a mechanism by which neurons exert top-down control across the brain (Cavanagh & Frank, 2014), including subcortical networks in the basal ganglia (Zink et al., 2006). These subcortical basal ganglia networks can be modulated by novelty; indeed, a human intraoperative study by Mikell and colleagues (2014) found that subcortical neurons in the substantia nigra increased firing more following infrequent novel sounds than frequent standard sounds.

Midfrontal delta/theta activity is attenuated in PD during attentional orienting, conflict, interval timing, and responses to startling stimuli (Cavanagh et al., 2018; K.-H. Chen et al., 2016; W. Chen et al., 2018; Güntekin et al., 2020; He et al., 2017; Kelley et al., 2018; Lange et al., 2016; Parker et al., 2015; Singh et al., 2018, 2021; Solís-Vivanco et al., 2018). Furthermore, during interval timing, midfrontal ∼4-Hz rhythms are coherent with field potentials in the subthalamic nucleus (STN) (Kelley et al., 2018). Notably, attenuated delta (1-4 Hz) activity during interval timing is predictive of cognition as measured by the Montreal Cognitive Assessment (MoCA), suggesting that the recruitment of midfrontal low-frequency oscillations reflects general cognitive-control processes (Singh et al., 2021). These data lead to the hypothesis that midfrontal ∼4 Hz rhythms engage subcortical networks in response to novelty as a part of an overall system of cognitive control.

In the current report, we tested this hypothesis by first comparing midfrontal EEG during an oddball task between individuals with PD and healthy older adult participants (Andrés et al., 2006; Courchesne et al., 1975; Escera et al., 1998; Parmentier et al., 2008, 2010). In addition, we recorded midfrontal EEG and intracranial STN neurons during an intraoperative oddball task in PD patients undergoing DBS implantation surgery. We report three main findings. First, we found that 1-4 Hz activity correlated with novelty-related slowing, whereas individuals with PD had decreased novelty-responsive midfrontal 4-7 Hz rhythms compared to controls. Second, we found that neurons in the STN were strongly response-modulated rather than cue-modulated, and most response-modulated STN neurons were influenced by novelty. Third, we found that midfrontal low-frequency 1-4 Hz oscillations were coherent with response-modulated STN neurons. These data provide insight into how novel information engages a midfrontal cognitive control system, and how this system in turn contributes to a frontal-subcortical mechanism for response control.

## MATERIALS AND METHODS

### Participants

This investigation included both an EEG experiment and an intraoperative neurophysiology experiment. For the EEG experiment, we recruited PD participants and demographically-similar control participants (see Table 1 for demographics and PD characteristics). PD participants were recruited through the Movement Disorders clinic at University of Iowa Hospitals and Clinics (UIHC). Healthy older adults were recruited to serve as control participants from either the Seniors Together in Aging Research (STAR) registry or a list of people who previously participated in research in our lab and agreed to be contacted for new research opportunities. Participants were considered healthy if they did not have any neurological and psychological diseases or disorders. Participants were recruited by email or phone and received compensation of $30/hour. All procedures were approved by the University of Iowa (UI) Institutional Review Board (IRB) (#201707828) and all participants provided informed consent.

**Table 1:** EEG participant demographics.

| Group | Gender | N | Age | Age | Right | Education | MoCA | UPDRS | LEDD | Disease |
| --- | --- | --- | --- | --- | --- | --- | --- | --- | --- | --- |
|  |  | (%) | median | range | Handed | median | median | Part 3 | median | Durati |
|  |  |  | yrs | yrs | (%) | yrs | (Q1-Q3) | median | (Q1 – Q3) | media |
|  |  |  | (Q1-Q3) |  |  | (Q1-Q3) |  | (Q1-Q3) |  | yrs |
|  |  |  |  |  |  |  |  |  |  | (Q1-Q |
| **Control** | Total | 35 | 71.0 | 52-86 | 32 | 16.0 | 27.0 | - | - | - |
|  |  |  | (64.0-75.5) |  | (91%) | (15.0-18.0) | (26.0-28.0) |  |  |  |
|  | F | 19 | 68.0 | 52-86 | 17 | 16.0 | 27.0 | - | - | - |
|  |  | (54.3%) | (62.5-72.5) |  | (89.4%) | (14.0-17.0) | (27.0-28.0) |  |  |  |
|  | M | 16 | 71.5 | 60-85 | 15 | 17.0 | 26.0 | - | - | - |
|  |  | (46.7%) | (67.0-78.0) |  | (93.8%) | (16.0-18.0) | (26.0-28.0) |  |  |  |
| **PD** | Total | 50 | 68.0 | 52-86 | 45 | 15.0 | 25.5 | 11.5 | 850 | 4.5 |
|  |  |  | (60.3-72.0) |  | (90%) | (13.0-18.0) | (22.3-27.0) | (7.0-17.0) | (462.5-1150.0) | (2.0-6. |
|  | F | 19 | 68.0 | 53-86 | 16 | 16.0 | 27.0 | 12.0 | 600 | 5.0 |
|  |  | (38.0%) | (66.0-71.5) |  | (84.2%) | (14.0-18.0) | (24.5-28.0) | (7.5-16.5) | (375.0-937.5) | (2.0-7. |
|  | M | 31 | 66.0 | 52-84 | 29 | 14.0 | 24.0 | 11.0 | 950 | 4.0 |
|  |  | (62.0%) | (60.0-73.0) |  | (93.5%) | (12.0-17.8) | (22.0-26.0) | (6.0-17.0) | (587.5-1315.0) | (2.5-6. |
| Control |  | p=0.21 | p=0.19 |  | p=1.0 | p=0.10 | p=0.001 |  |  |  |
| vs PD |  | (M vs | (Wilcoxon) |  | (R vs L, | (Wilcoxon) | (Wilcoxon) |  |  |  |
| | | F, $\chi^2$) | | | $\chi^2$) | | | | | |

All participants in the EEG experiment were tested on levodopa as our past work has shown that levodopa does not reliably modulate midfrontal ∼4 Hz rhythms (Singh et al., 2018, 2021). We performed the Unified Parkinson’s Disease Rating Scale (UPDRS) Part 3 during our experimental sessions while participants were on dopaminerigic medication to measure symptom severity nearest the time of task completion.

In parallel with the EEG experiment, we recruited 18 PD patient-volunteers who had elected to undergo bilateral STN deep brain stimulation (DBS) implantation surgery (see Table 2). These participants were recruited over a time period of 1.5 years (July 2019–August 2020). Within the study period there was a 5-month pause in enrollment due to research suspension during the COVID-19 crisis. Patients were asked during preoperative clinical sessions (1–14 days prior to surgery) whether they were interested in participating in research during their DBS implantation surgery. If they agreed, a researcher separate from the surgical team discussed the research protocols with them, obtained informed consent, and completed a brief practice session of the task. Participant demographics and PD characteristics are presented in Table 2. Note that these participants in the intraoperative study were tested off PD medications, as is necessary for the clinical placement of the deep brain electrodes. All intraoperative research procedures were approved by the UI IRB (#201402730).

**Table 2.** Intraoperative patient-volunteer demographics.

| **Gender** | **N** | **Age** | **Age range** | **Right** | **LEDD** | **Disease** |
| --- | --- | --- | --- | --- | --- | --- |
|  |  | **median** | **yrs** | **Handed** | **median** | **Duration** |
|  |  | **yrs** |  | **(%)** | **(Q1-Q3)** | **median** |
|  |  | **(Q1-Q3)** |  |  |  | **yrs** |
|  |  |  |  |  |  | **(Q1-Q3)** |
| **Total** | 18 | 67.0 | 53-71 | 15 (83%) | 975.0 | 7.0 |
|  |  | (62.3-68.5) |  |  | (687.5-1175.0) | (3.5-10.8) |
| **F** | 4 (22%) | 62.5 | 53-71 | 3 | 921.5 | 12.0 |
|  |  | (57.5-68.0) |  | (75%) | (819.8-987.5) | (10.8-13.5) |
| **M** | 14 (78%) | 67.0 | 53-71 | 12 (86%) | 1018.5 | 6.5 |
|  |  | (63.0-68.5) |  |  | (687.5-1218.8) | (3.0-7.0) |

### Oddball task for EEG experiment

We assessed the response to novelty using a version of the cross-modal oddball distractor task (Andrés et al., 2006; Escera et al., 1998; Parmentier et al., 2008, 2010; Parmentier & Hebrero, 2013; Wessel & Huber, 2019). This task was presented using the PsychToolbox-3 functions (Brainard, 1997; Kleiner et al., 2007; Pelli, 1997, http://psychtoolbox.org/) in MATLAB 2018b on either a Linux or Windows computer. Task-specific audio was played through Dell Rev A01 speakers positioned on either side of the monitor. Responses were made with the left and right index fingers on a standard QWERTY USB-keyboard. In addition to the cross-modal oddball distractor task, participants completed additional cognitive and motor tasks, as well as detailed neuropsychological and neuropsychiatric assessments as part of other protocols (Singh et al., 2018, 2021).

Our version of the cross-modal oddball distractor task involved a choice reaction-time task in which a white arrow appeared in the center of a black screen, and the participant was required to press the key that corresponded with the direction of the arrow (“q” for left arrow, “p” for right arrow) as quickly as possible. The appearance of the arrow was preceded by an audio-visual cue by 500 ms (either the standard cue or the distractor cue). Participants were instructed that this cue would appear 500 ms before the target stimulus (white arrow), and they were told that the cue would be a green circle and a short tone (600-Hz sine wave tone lasting 200 ms). The audio-visual cue was followed by the target arrow (**Figure 1A**). Participants had to respond within 1 s, after which the fixation cross reappeared and the next trial started after a variable inter-trial interval between 500–1000 ms.

**Figure 1.**
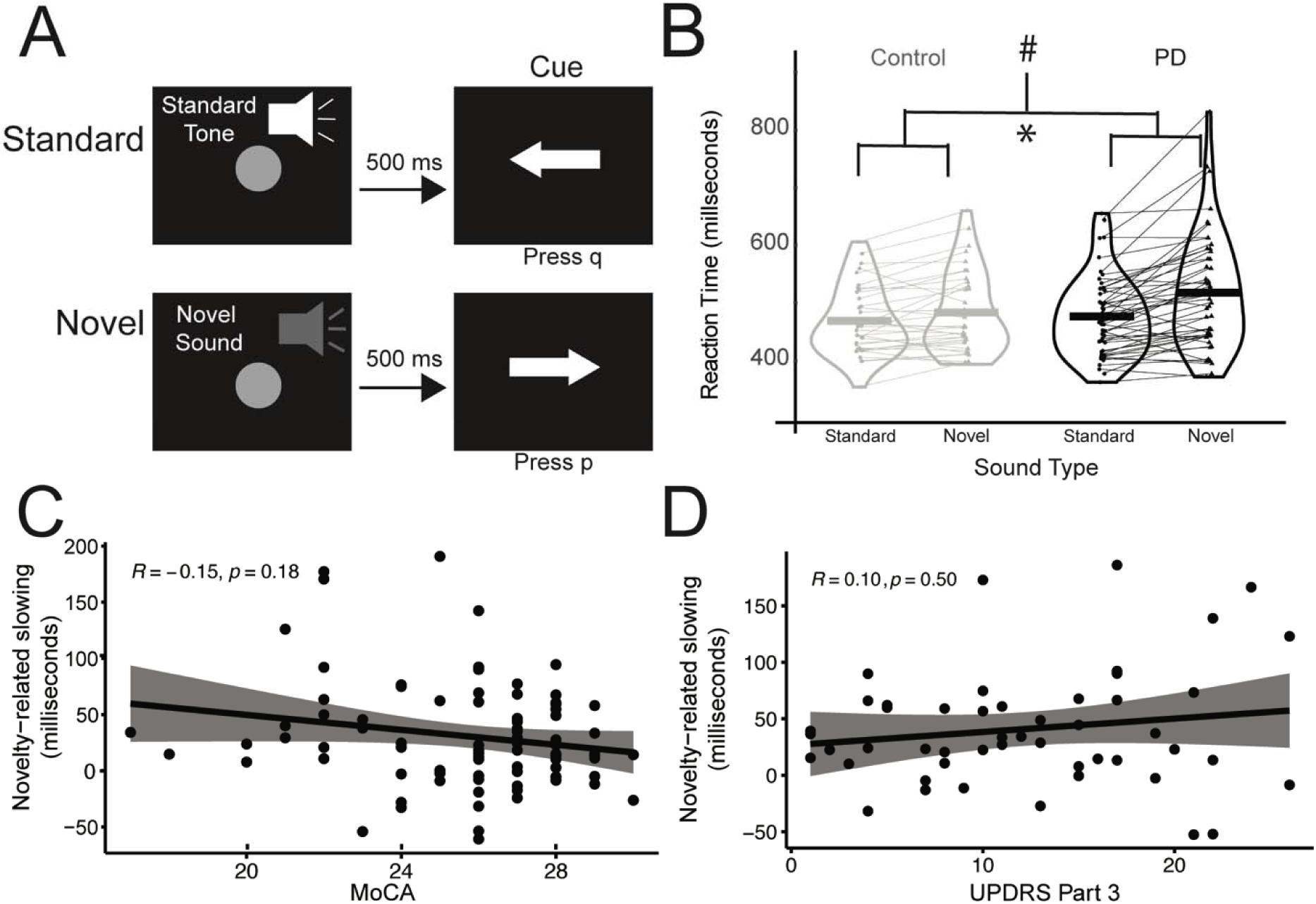
Novelty slows responses more for PD participants than controls. A) The cross-modal oddball distractor task consisted of trials with an arrow preceded by either a standard tone (80% of trials) or a novel sound (10%; of note, 10% of trials used a novel visual stimulus; these trials were not analyzed). The arrow required a left “p” or right “q” key press, based on the direction. B) Reaction time for Control and PD participants; each dot represents an individual’s median RT and the thick horizontal bar represents group medians. Novel oddball stimuli disproportionately distracted PD participants. Compared to Control participants, PD participants responded more slowly to the arrow on trials when a novel sound was presented relative to a standard tone. * indicates a significant main effect of sound type (standard vs. novel) on reaction time. # indicates a significant interaction between sound type and group (control vs. PD) on reaction time via a linear mixed-effects model. C) Cognitive function as measured by the Montreal Cognitive Assessment (MoCA) did not significantly correlate with novelty-related slowing. D) UPDRS Part 3 did not significantly correlate with novelty-related slowing in PD participants.

Participants completed a brief practice of the task (10 trials), which had all the familiar standard cues described above. Following the practice, participants completed four blocks of 60 trials each, for a total of 240 trials. Of the 240 trials, 80% contained the standard cues as described above, 10% contained an unexpected auditory oddball cue (non-repeating, randomly-created sine wave that sounded like a birdcall or robotic noise lasting 200 ms in duration) in place of the expected tone, and 10% contained an unexpected visual oddball cue (unique shape/color combination) in place of the green circle. Because visual stimuli were not presented during intraoperative research, only auditory oddball trials were analyzed in both datasets.

### EEG recordings

EEG recordings were performed according to methods described in detail previously (Singh et al., 2020, 2021). Briefly, we used a 64-channel EEG actiCAP (Brain Products GmbH) with a 0.1-Hz high-pass filter and a sampling frequency of 500 Hz. We used electrode Pz as reference and electrode FPz as the ground. EEG activity was referenced according to the procedures described in Singh et al., (2020, 2021). An additional channel was recorded at the mid-inion region (Iz), and we removed unreliable Fp1, Fp2, FT9, FT10, TP9, and TP10 channels, resulting in 59 channels for pre- and post-processing. Data were epoched around the cues from -1000 ms to 2500 ms peri-cue.

Bad channels and bad epochs were identified using the FASTER algorithm (Nolan et al., 2010) and the pop_rejchan function from EEGLAB, and were then interpolated and rejected respectively. On average, 1.6 ± 0.9 channels per subject were removed, and Cz was never removed during preprocessing. Eye blink contaminants were removed following independent component analysis (ICA).

Event-related potentials (ERPs) were low-pass-filtered at 20 Hz for analyses. We calculated average ERPs for each trial type and each group (PD and control). Our primary interest was in ∼4-Hz midfrontal rhythms; consequently, we utilized time-frequency analyses. After preprocessing, there was no additional filtering for this analysis. We computed spectral measures by multiplying the fast Fourier transformed (FFT) power spectrum of single-trial EEG data with the FFT power spectrum of a set of complex Morlet wavelets (defined as a Gaussian-windowed complex sine wave: e^i2πtf^e^-t^2/(2xσ ^2)^, where *t*=time and *f*=frequency). Wavelets increased from 1– 50 Hz in 50 logarithmically-spaced steps, which defined the width of each frequency band, increasing from 3–10 cycles between 1–50 Hz and taking the inverse FFT (Cohen, 2014). The end result of this process was identical to time-domain signal convolution, and it resulted in estimates of instantaneous power (the squared magnitude of the analytic signal) and phase angle (the arctangent of the analytic signal). We then cut the length of each signal accordingly for each trial (−500 ms to 1000 ms). These short temporal epochs reflect the wavelet-weighted influence of longer time and frequency periods. Power was normalized by converting to a decibel (dB) scale (10*log10(power_t_/power_baseline_)), allowing us to directly compare the effects across frequency bands. The baseline for each frequency was calculated by averaging power from □300 ms to -200 ms prior to cue onset (Singh et al., 2018, 2020, 2021). A 100 ms duration is often used as an effective baseline since pixel-wise time-frequency data points have already been resolved over smoothed temporal and frequency dimensions with the wavelets. We took an *a priori* approach focused on channel Cz and delta and theta-band tf-ROIs (300-400ms, delta: 1-4 Hz and theta: 4-7 Hz); these tf-ROIs are strongly justified based on our extensive past work in cognitive control tasks, EEG, and PD (K.-H. Chen et al., 2016; Kelley et al., 2018; Parker et al., 2015; Singh et al., 2018, 2020, 2021). Technically, due to the log-scale of the frequency bands, our delta ROI was defined by the frequency bins 1.0233 - 3.9443, and our theta ROI was defined by the frequency bins 4.2701 – 7.442.

### Oddball task for intraoperative neurophysiology

We used an auditory version of a 3-tone oddball task during intraoperative human neurophysiology to capture neuronal novelty during DBS-electrode implantation surgeries. Task design was driven by clinical requirements and the physical experimental set-up in the operating room. This task is ideal for research for intraoperative patients whose heads are in a stereotaxic frame during multielectrode recordings because 1) it is in the auditory domain and does not require a monitor for visual stimuli, 2) it is relatively simple in that it requires a motor response to sounds, and 3) several trials can be collected in a few minutes. Similar 3-tone designs have been used previously (Debener et al., 2005), in which novelty is probed with novel tones. All stimuli were auditory and presented at an appropriate volume through earbuds or external speaker to be clearly heard over operating room noise. The task and stimuli were presented using PsychToolbox-3 in MATLAB 2018b on a Windows laptop. Responses were made with a Kinesis pedal that the patients held in their hands. Participants pressed buttons with their thumbs to respond.

For the 3-tone oddball task, each trial consisted of one of the following sounds being played: a standard tone that occurred frequently (500 Hz: 50% of trials), a target sound that occurred infrequently (the word “Go”: 30% of trials; this trial type was used to maintain attention), or a novel sound (unique birdcall: 20% of trials; **Figure 4A**). Patients were instructed to press with their left hand (typically thumb) if they heard the word “Go,” and to press with their right hand/thumb if they heard anything else. We focused our analyses on the standard and the novel tones, as patients responded with their right hand for both the standard tones and the novel oddball tones, facilitating a direct comparison of novelty induced motor initiation. Patients practiced the task with 30 trials preoperatively and again on the morning of surgery. The intraoperative experiment was conducted after the clinically-necessary microelectrode recording electrodes were confirmed to be in the STN based on clinical recordings.

### Intraoperative neurophysiology recordings

Bilateral DBS electrodes were implanted sequentially during a single stereotactic procedure. Participants received short-acting pain relief and sedative medications such as remifentanil and/or dexmedetomidine; these were stopped >1 hour prior to the task for participants to be maximally awake for necessary clinical testing and participation in research. Patients underwent standard bilateral DBS lead implantations using indirect framed stereotactic STN targeting, refined by microelectrode recordings from 0.4 to 0.8 mΩ tungsten electrodes (Alpha-Omega, Inc). Three simultaneous microelectrode recording tracks were used, consisting of anterior, middle, and posterior trajectories, separated by 2 mm center-to-center from an entry point near the coronal suture. STN margins were defined by the functional and electrical properties from these microelectrode recordings, in line with standard clinical practice. Because microelectrode recordings were clinically necessary, participants were not exposed to extra electrode penetrations to participate in the study.

In addition to STN recordings, scalp EEG was also simultaneously recorded in the operating room during the oddball task. Prior to the start of surgery, three midfrontal EEG electrodes (Cadwell Industries, Inc) were placed over frontal locations on the forehead at the hairline with right and left electrodes approximating F1 and F2 and midfrontal lead approximating Fz (**Figure 5A**). A reference electrode was placed on the left mastoid. We recorded EEG because this location indexes midfrontal ∼4-Hz rhythms and is in line with prior work (K.-H. Chen et al., 2016; Kelley et al., 2018; Narayanan et al., 2013; Singh et al., 2021). EEG recordings were recorded at 24 kHz and subsequently downsampled to 500 Hz. To adjust for the noisy recording environment of the operating room, the signal was filtered below 50 Hz with a low-pass filter, followed by a 1-Hz high-pass filter. Microelectrode recordings were sampled at 24 kHz, amplified, and filtered for single neurons (1–8 kHz) and STN local field potentials (<200 Hz). Neurophysiological and behavioral data were acquired simultaneously using a Tucker-Davis Technologies multi-channel data acquisition system.

After the experiments, Offline Sorter (Plexon) was used to analyze STN activity and remove artifacts. Spike activity was analyzed for all cells that fired at rates above 0.1 Hz. Principal component analysis (PCA) and waveform shape were used for spike sorting. Single units were defined as those 1) having a consistent waveform shape, 2) being a separable cluster in PCA space, and 3) having a consistent refractory period of at least 2 ms in interspike interval histograms. Isolated STN single units were included for analyses if 1) the recording location was confirmed by the neurosurgeon to be in the STN based on clinically-necessary and observable STN firing patterns during surgery, and 2) the unit was held throughout the ∼6 minute duration of the task.

### Experimental Design and Statistical Analyses

All data and statistical approaches were reviewed by the UI Biostatistics, Epidemiology, and Research Design Core at the Institute for Clinical and Translational Science. All code and data are available at narayanan.lab.uiowa.edu. For demographic summary statistics, medians and interquartile ranges were calculated for continuous measures. Counts and percentages were calculated for categorical measures. EEG participant demographics were stratified by group and gender. Intraoperative participant demographics were stratified by gender. We used the nonparametric Wilcoxon rank sum test to test for differences in age, education, and MoCA between strata. We also used a chi-squared test to test for differences in proportion of male/female and handedness between groups. For behavioral data from the EEG study, we used a linear mixed-effects model with random intercept for participants’ response time to capture the effects of predictor variables (tone type: standard tones vs novel sounds and group: PD vs control) on response time. For behavioral data for the intraoperative study, we used the nonparametric Wilcoxon rank sum test to test for differences between sound stimuli on accuracy and reaction time. For EEG data, we used linear mixed-effects models with random intercept for participants’ time-frequency power to compare average time-frequency power between groups. The fixed-effect predictors included group, tone type, MoCA score, and UPDRS Part 3 as control variables and time-frequency power from the *a priori* regions of interest (ROIs) as the outcome variables. We set our alpha for these analyses at 0.05. The MoCA score was included in the model as a covariate given that it was different between groups. UPDRS Part 3 was included as a covariate for PD participants in order to account for degree of motor impairment. These analyses were conducted in R.

Neuronal modulations were defined via a generalized linear model (GLM) for each neuron, where the outcome variable was firing rate, and the predictor variable was cue or response, consistent with past work (Emmons et al., 2019, 2020). For each neuron, we constructed a model in which the outcome variable was the firing rate binned at 0.1 seconds, and the predictor variable was either cue or response. Main effects comparisons were made stratified by neuron. Across the ensemble, *p* values were corrected via Benjamini-Hochberg false-discovery-rate (FDR; Benjamini & Yekutieli, 2001), with *p* values <0.05 considered significant modulation for each neuron. We also analyzed neuronal patterns using PCA of peri-event histograms around response binned at 0.1 second with kernel-density estimates (bandwidth 0.25). Finally, we analyzed midfrontal EEG-STN coherence with the Neurospec 2.0 coherence toolbox (https://www.neurospec.org/) using the sp2a_m1 function with a segment power of 9 and a window size of 250 ms (Rosenberg et al., 1989). We have used this toolbox extensively in the past to examine spike-field coherence within and across brain networks (Kelley et al., 2018; Kingyon et al., 2015; Narayanan et al., 2013; Parker et al., 2014). We used delta and theta ROIs comparable to the EEG study (delta: 1-4 Hz, theta 4-7 Hz) to test whether delta/theta-band activity was coherent with spiking activity in response-modulated and non-response-modulated neurons. To compare coherence across neurons with different spike rates and trial numbers, spike-field coherence was scaled such that the raw spike-field coherence of each neuron was divided by the 95% confidence interval for each neuron. According to this metric, a “1” represents coherence at the 95% confidence interval. These analyses were performed in MATLAB.

## RESULTS

### Demographics

Table 1 shows demographics for the EEG study and Table 2 shows demographics for the intraoperative study. Analysis for the EEG study was conducted with 50 individuals with PD and 35 control participants. We collected data from similar numbers in each group, but we experienced technical issues that caused the exclusion of data for a subset of participants. For EEG participants, demographic results were stratified by group and gender. Wilcoxon test results showed that the groups were not significantly different in terms of age (*p*=0.19, effect size r=0.14) or years of education (*p*=0.10, effect size r=0.18). The MoCA score was higher for the control participants compared to PD participants (*p*<0.001, effect size r=0.38), thus we accounted for MoCA in our analyses.

### PD participants had slowed reaction-times during novelty-related distraction

We studied how novelty engaged cortical cognitive control mechanisms by recording EEG during the cross-modal oddball distractor task (**Figure 1**). This task asked participants to respond to a target cue (arrow) following either a familiar stimulus (600-Hz tone) or a novel stimulus consisting of complex and unique auditory features.

Response accuracy was high overall (Median (1^st^ Quartile–3^rd^ Quartile); Control: standard 99.5% (99.5%-100.0%), novel 100.0% (100.0%-100.0%); PD: standard 98.2% (93.4%-99.5%), novel 97.9% (91.7%-100%)), although control participants were significantly more accurate than PD participants (main effect of group: *F*_(1,83)_=15.3, *p<*0.001). There was no main effect of sound type on accuracy (*F*_(1,83)_=0.52, *p*=0.47), and no interaction between group and sound type on accuracy (*F*_(1,83)_=2.3, *p=*0.13).

Regarding response speed, our model revealed a significant interaction between the effects of group and sound type on reaction time (*F*_(1,83)_=7.3, *p<*0.01), such that PD participants were disproportionately more affected by sound type than control participants. On trials with a distracting novel sound, control participants responded approximately as quickly as on trials with a standard tone (Median (1^st^ Quartile–3^rd^ Quartile); Control: novel 454 ms (427-527), standard 454 ms (416-508 ms)), whereas PD participants responded more slowly compared to trials with a standard tone (PD: novel 505 ms (439-571 ms), standard 462 ms (416-515 ms); **Figure 1B**).

There was a significant main effect of sound type (*F*_(1,83)_=31.7 *p*<0.001), which appears to be driven by the effect within the PD group. There was no significant effect of group (Control vs. PD) on reaction time (F_(1,83)_=1.7, *p*=0.20). Post-hoc analyses revealed that within the PD group, there was a significant difference between reaction times for the sound types (t(83.0)=-6.5, p<0.0001). There was also a significant difference in reaction time between controls on standard tones and PD participants on novel sounds (t(98.1)=-2.8, p=0.03). No other post hoc contrast was significant. As expected, controls and PD participants both showed significant effects of block on novelty-related slowing (control: F_(1,102)_=6.3, *p*=0.01; PD: F_(1,147)_=19.9, *p*<0.001). Both groups had greater novelty-related slowing in Block 1 compared to Blocks 2, 3, and 4.

Finally, we found that novelty-related slowing was not related to cognitive function, as measured by MoCA (Spearman ρ=-0.15, *p*=0.18; **Figure 1C**), or motor function, as measured by UPDRS Part 3 in PD participants (Spearman ρ=0.10, *p*=0.50; **Figure 1D**). Taken together, these data suggest that PD participants were more distracted by novel stimuli than controls.

### PD participants had attenuated ∼4-Hz midfrontal rhythms

Next, we examined cortical correlates of cognitive control in response to novelty. We found that ERPs were differentiated between controls and PD participants for standard tone trials (**Figure 2A, top panel**) and novel sound trials (**Figure 2A**, bottom panel). The possibility of an underlying low-frequency contribution across the entire ERP in PD participants suggests that time-frequency analysis might be more effective in isolating unique temporal events that differ between groups.

**Figure 2.**
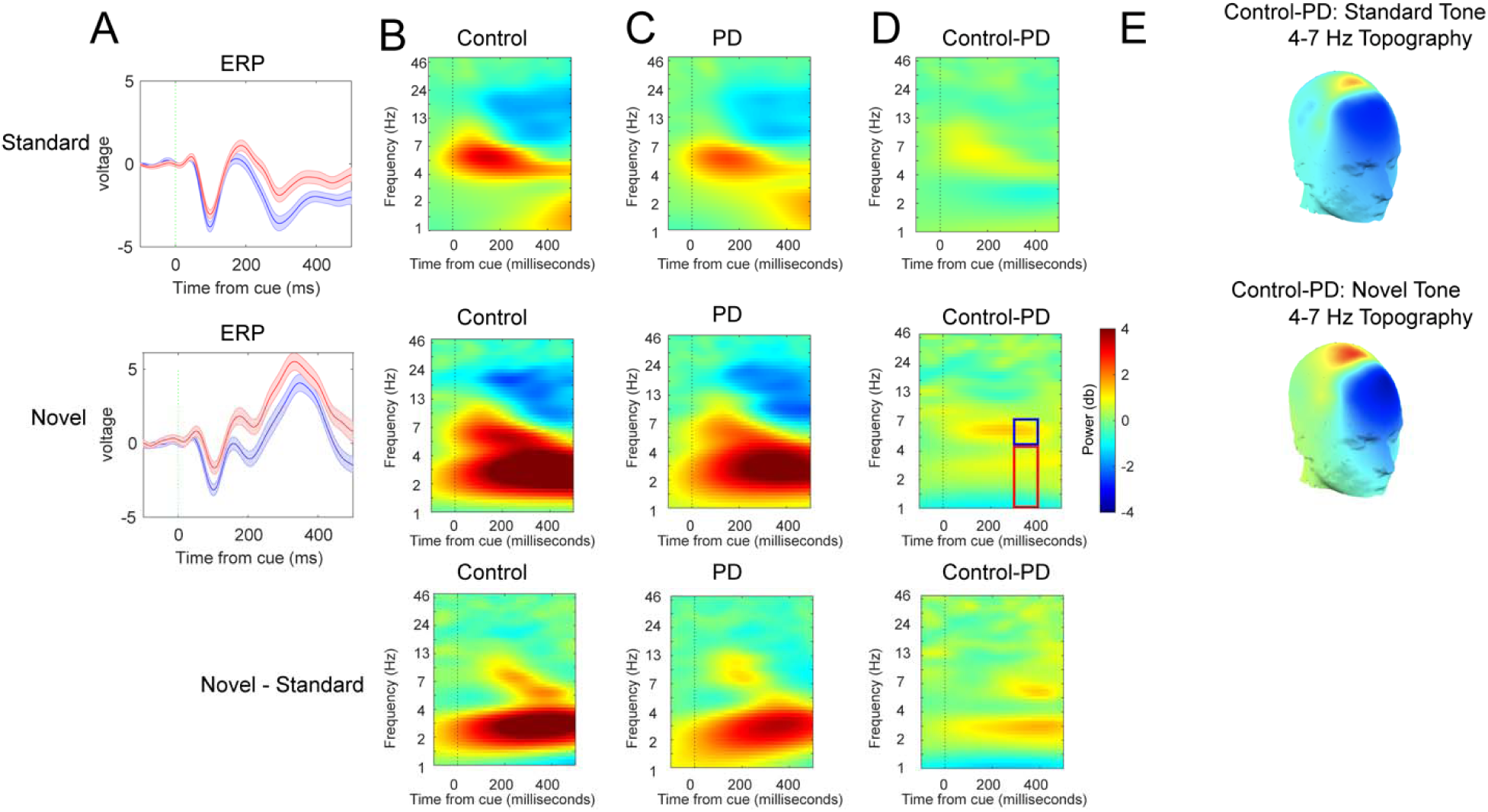
Novelty boosts midfrontal ∼4-Hz power, which is attenuated in PD. A) Average event-related potentials for trials with standard tones (top row) and novel tones (bottom row) for controls (blue line) and PD participants (red line). B) Time-frequency power spectrograms for standard tones (top panels), novel sounds (middle panels), and novel – standard (bottom panels) for Control and C) PD, and D) Subtraction of Control – PD. The red box in D (middle panel) represents the delta band region-of-interest (ROI) (1-4), while the blue box represents the theta band ROI (4-7 Hz) 300-400 ms after the cue. E) Topographical representation of the comparison of theta power between controls and PD participants (Control – PD) for standard tones (top panel) and novel sounds (middle panel). Data from 50 PD and 35 control participants.

We examined power from *a priori* tf-ROIs (delta: 1-4 Hz and theta: 4-7 Hz for 300-400 ms post-tones; blue and red boxes in **Figure 2D**). For our delta ROI, we found no interaction between group and sound type (F(1,83)=0.88,*p*=0.35) and no main effect of group (F(1,81)=0.02, *p*=0.89; **Figure 3A**). Our linear mixed-effects model did reveal a significant effect of sound type (F(1,83)=166.29, *p*<0.0001), such that delta power was higher for novel sound trials compared to standard tone trials. There were no significant effects of MOCA or UPDRS Part 3. For theta, there was a significant interaction between sound type and group (*F*(1,83)=6.6, *p=*0.01; **Figure 3B**), such that controls had higher power in this ROI during trials with a novel sound compared to a standard tone, whereas PD participants did not have as large of a difference between sound types. Similar to delta, our model revealed a significant main effect of sound type on this tf-ROI (*F*(1,83)=47.91, *p*<0.001), with novel sound trials showing higher theta power. There were no significant main effects of group (F(1,81)=0.61,*p*=0.44), MoCA, or UPDRS Part 3.

**Figure 3.**
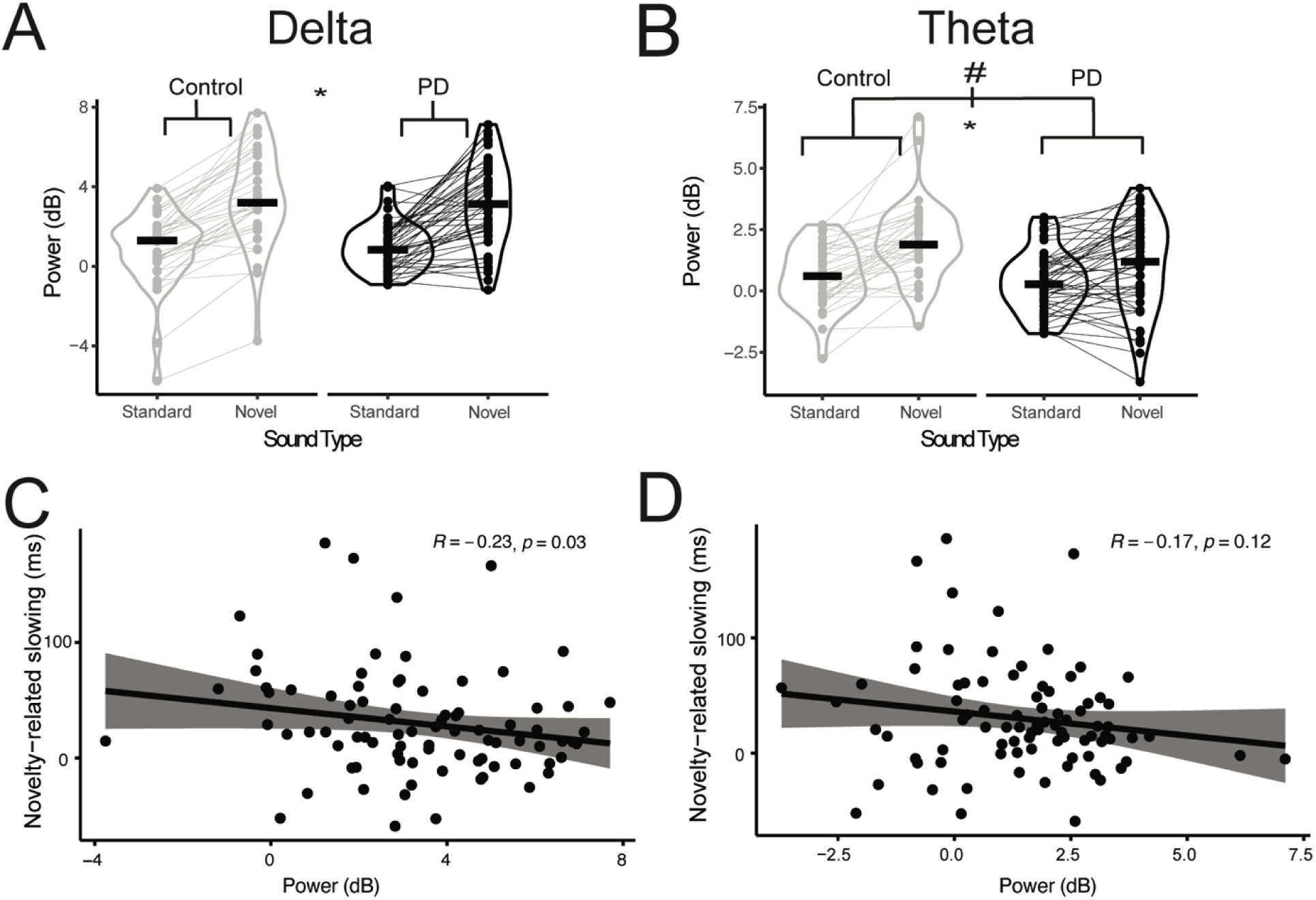
1-4 Hz delta rhythms are linked with novelty-related slowing, and 4-7 Hz theta rhythms are attenuated in PD. A) Power in a priori delta tf-ROI (1-4 Hz; 300-400 ms after cue) for Control and PD participants; each dot represents an individual’s mean power and the thick horizontal bars represent group medians. A linear mixed-effects model revealed a significant main effect of sound type (novel vs. standard), denoted by *. B) Power in a priori theta tf-ROI (4-7 Hz; 300-400 ms after cue) for Control and PD participants. A linear mixed-effects model revealed a significant interaction between sound type and group on power in this ROI, denoted by #, and a significant main effect of sound type (novel vs. standard), denoted by *. Compared to control participants, PD participants experienced a smaller increase in theta power on trials when a novel sound was presented relative to a standard tone. C) For the delta tf-ROI, there was a significant relationship between novelty-related slowing and delta power across all participants; but D) for the theta tf-ROI, this relationship did not achieve significance. Data from 85 participants (35 Control and 50 PD).

We found a significant relationship between midfrontal 1-4 Hz power and novelty-related slowing (Spearman ρ*=*-0.23, *p=*0.03; **Figure 3C**). However this relationship with behavior was not significant for midfrontal 4-7 Hz power (Spearman ρ*=*-0.17, *p=*0.12; **Figure 3D**). Finally, delta power on novel trials did not correlate significantly with MOCA (Spearman ρ=0.08, *p*=0.43) or UPDRS Part 3 (for PD group only; Spearman ρ*=*-0.04, *p*=0.78). Similary, 4-7 Hz power also did not correlate significantly with MoCA (Spearman ρ*=*0.12, *p*=0.28) or UPDRS Part 3 (for PD group only; Spearman ρ*=*-0.23, *p*=0.11). These data are convergent with past work showing that 1-4 Hz rhythms are linked with response control (Singh et al., 2021) whereas 4-7 Hz rhythms are attenuated in PD (Güntekin et al., 2020; He et al., 2017; Iyer et al., 2020; Kelley et al., 2018; Parker et al., 2015; Singh et al., 2018, 2021; Soikkeli et al., 1991).

### Human single STN neurons were modulated around responses to novelty

To investigate the cortico-striatal pathways underling novelty-modulated responses, we leveraged intraoperative neurophysiology to record from single STN neurons. Participants performed the task with a median accuracy of 87% (Q1-Q3: 82%-92%); there was no difference in median accuracy or reaction time between tone types (accuracy: standard 86% (77%-93%), novel 90% (80%-95%), *p*=0.21, effect size r=0.71; reaction time: standard 877 ms (721-1024 ms), novel 875 ms (761-936 ms), *p*=0.99, effect size r=0.71; **Figure 4B**).

**Figure 4.**
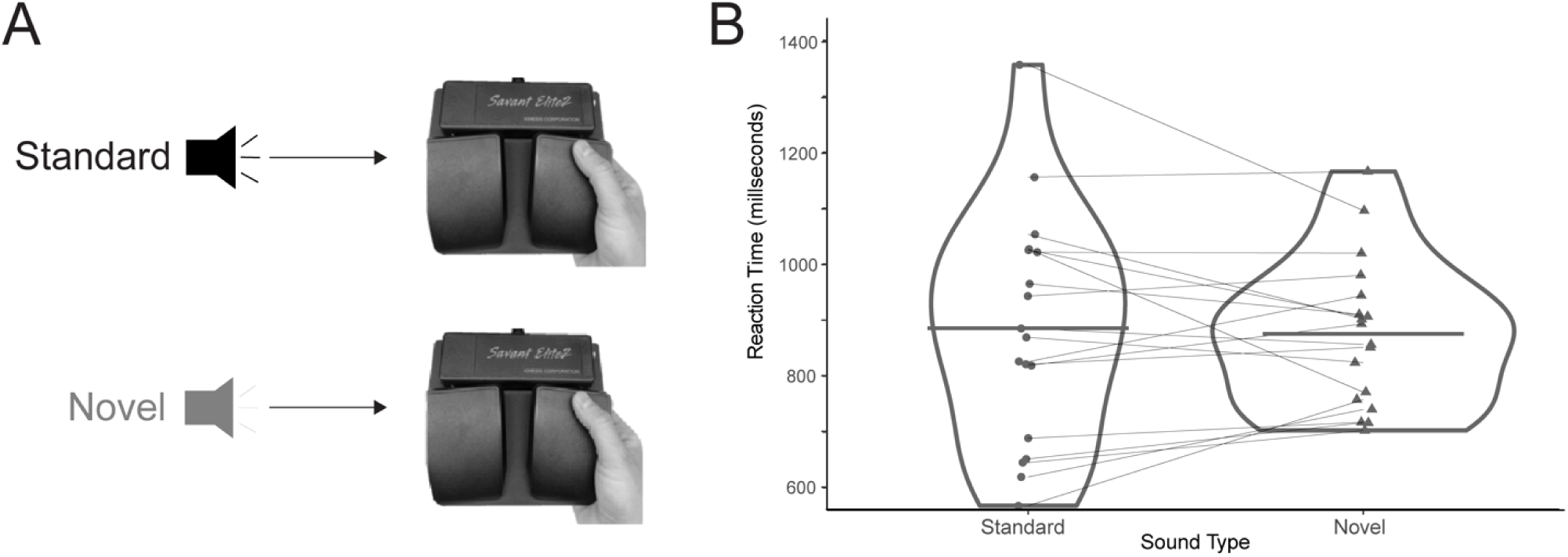
Intraoperative recordings of novelty-related responses. A) Auditory 3-tone oddball task performed by patients with PD in operating room during deep brain stimulation surgery. Patients responded to standard tones (50% of trials) and novel sounds (20% of trials) with a right-hand response using a hand-held Kinesis pedal; an additional trial type with a constant stimulus required a left response and was not analyzed. B) Reaction times for standard tones and novel oddball sounds; each dot represents individual median reaction times, and thicker horizontal lines represent group median reaction times. No significant differences for reaction times between groups were found. Data from 18 PD patient-volunteers during intraoperative neurophysiology.

With these 18 participants, we also performed intraoperative scalp EEG (**Figure 5A**). Although these recording locations were anterior to the midfrontal areas which showed peak 1-7 Hz activity in the EEG experiment outside the operating room (i.e. **Figure 2)**, we were able to capture midfrontal cue-related activity from the intraoperative participants (**Figure 5B-C**).

**Figure 5.**
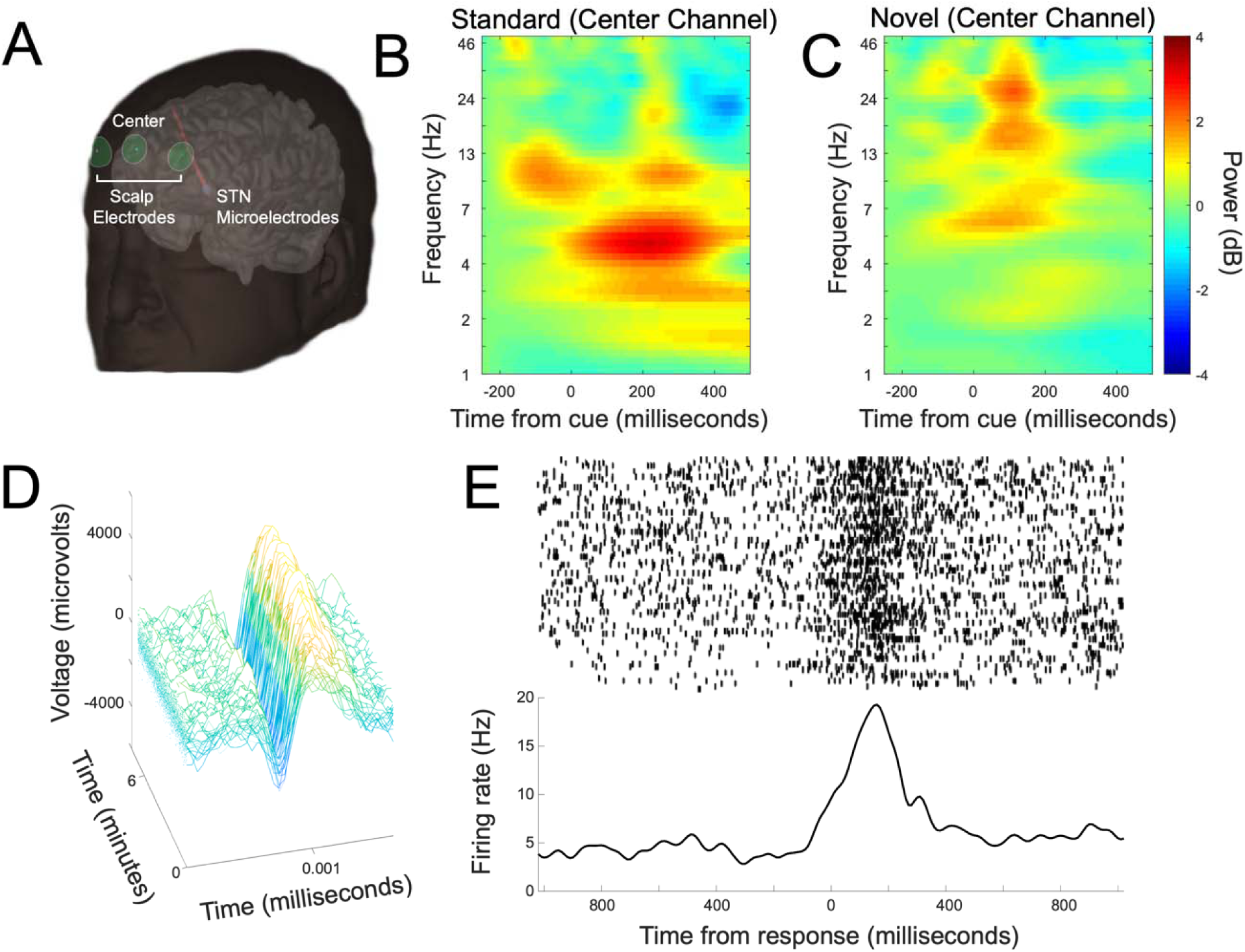
Recording simultaneously from frontal lobe and STN during surgery. A) Schematic of surgical recording with three frontal scalp-based electrodes (center, right, and left channels), and STN microelectrode recordings. B) Time-frequency plots of activity from the center channel for standard and novel tones in the auditory 3-tone oddball task. Data from 18 PD patient-volunteers undergoing DBS implantation surgery. D) Waveforms from a single STN neuron indicating that neuronal waveforms of voltage vs. time were stable over ∼6 minutes. E) Response-locked peri-event raster of neuronal firing from an exemplar neuron; the top panel is a peri-response raster, with each row representing one trial and each tick representing an action potential. The bottom panel represents a peri-event histogram of firing rate around all responses.

We identified 54 well-isolated STN neurons from 17 of the 18 participants (one did not have STN neuronal recordings; **Figure 5D-E**). We defined modulations via neuron-by-neuron GLMs of firing rate vs. all cues and responses at a trial-by-trial level. We found that 5 of the 54 neurons (9%) were significantly modulated around the cue. By contrast, we found significantly more neurons (17 of 54, 32%) with response-related activity (*χ*^2^=8.2; p=0.004; **Figure 5E & Figure 6A–B**). To further characterize the patterns of neuronal activity, we utilized PCA, a proven data-driven technique, to characterize neuronal activity (Chapin & Nicolelis, 1999; Narayanan & Laubach, 2009). We found that the first component (principal component 1; PC1) explained 38.5% of variance and appeared to be modulated around response; PC2 explained 28.5% of variance and was modulated prior to response (**Figure 6C-D**). These results provide evidence that STN neurons were strongly response modulated.

**Figure 6.**
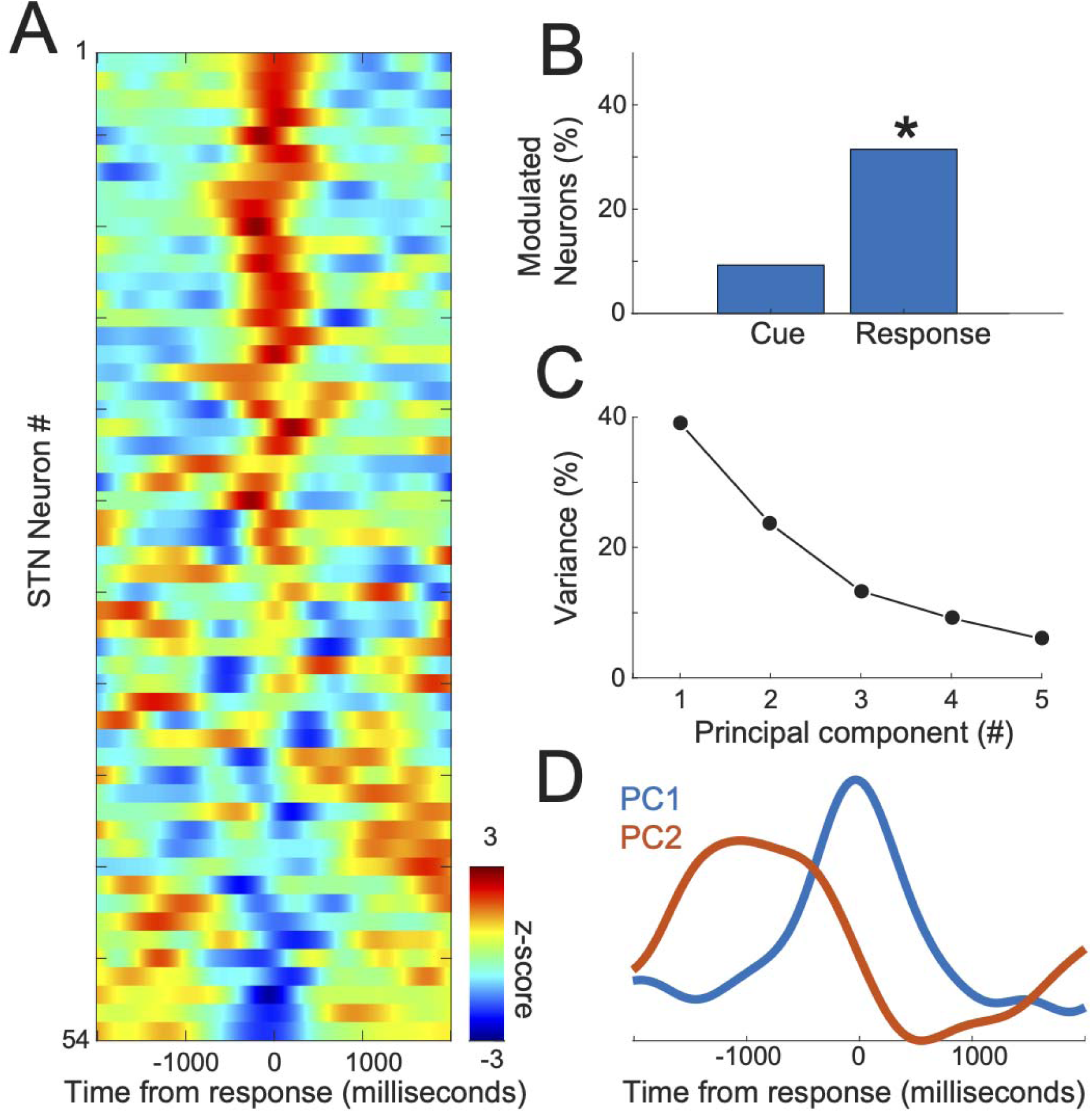
STN neurons had prominent response modulations. A) Average z-scored activity of all 54 STN neurons around responses. B) Trial-by-trial generalized linear models of firing rate vs. cue and response revealed that STN neurons were more strongly modulated by response than cue; *indicates significance via a *χ*^2^ test. C) Principal component analysis (PCA), a data-driven approach to identify neuronal patterns across an ensemble, revealed that principal components 1 and 2 (PC1 and PC2) explained 64% of variance among STN ensembles. D) PC1 and PC2 were modulated around response. Data from 54 STN neurons in 17 PD patient-volunteers.

Next, we were interested in how response-related activity was modulated by novel cues. Strikingly, we found that nearly all (16 of 17; 94%; **Figure 7B**) response-related neurons had differential activity with novel cues as quantified by an FDR-corrected neuron-by-neuron GLM of event type on response-related activity. Thus, these data indicate that nearly all STN response-related activity was modulated by novelty.

**Figure 7.**
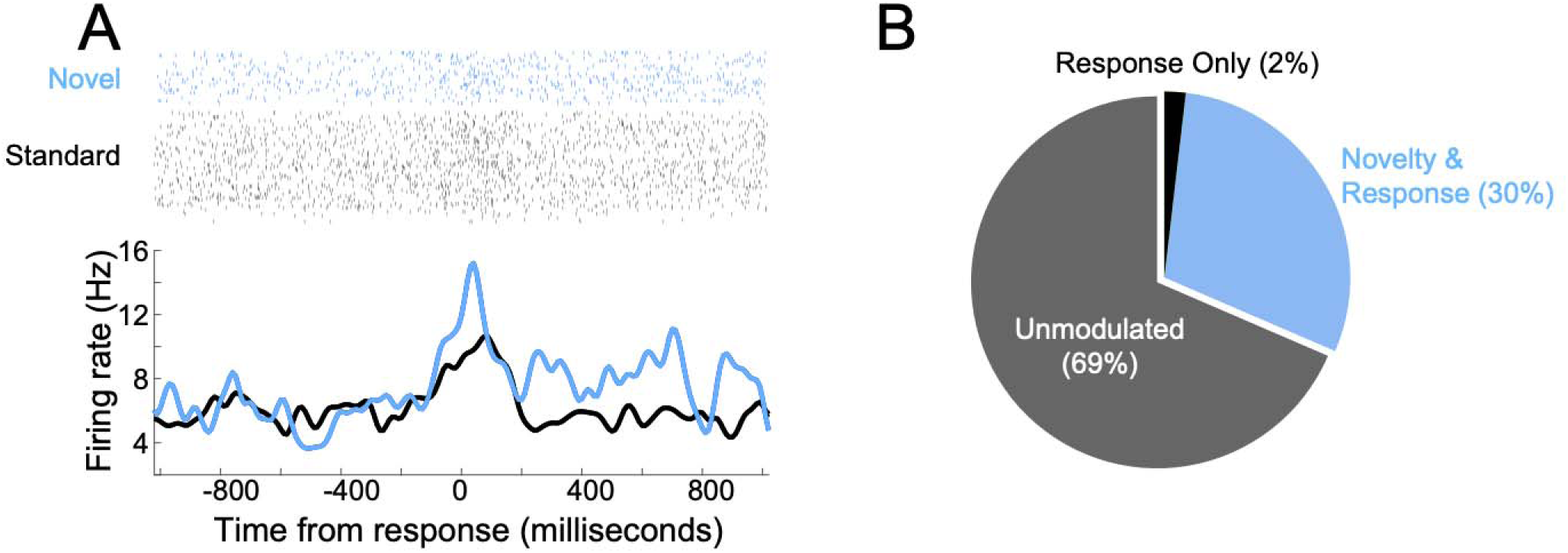
Most response-related neurons in the subthalamic nucleus were modulated by novelty. A) Example raster of neuronal spiking from one novelty-modulated neuron. B) We recorded from 54 STN neurons. Of these, 17 STN neurons had response-modulations. 16 were modulated by novelty and response (30% of the total), and 1 was modulated by response only (2% of the total). Data recorded in 17 patient-volunteers.

### Response-related STN neurons showed low-frequency coherence with midfrontal EEG

Finally, we were interested in the relationship between STN neuronal activity with scalp EEG-related modulations. We examined spike-field coherence, through which we could link the activity of midfrontal scalp electrodes to STN spiking (Kim & Narayanan, 2019; Narayanan et al., 2013; Parker et al., 2014, 2015). For some STN neurons, we noticed that spikes fired in-phase with low-frequency scalp EEG activity (**Figure 8A**). Among response-related STN neurons, there was significant low-frequency spike-field scalp-STN coherence after the cue around the time of responses (**Figure 8B**). Around responses, low-frequency delta coherence was stronger for response-related neurons compared to non-response-related neurons (**Figure 8C vs 8B**; scaled spike-field coherence for response neurons: 0.6(0.4-1.5) vs. non-response neurons 0.3(0.2-0.6); Wilcoxon rank sum *p*=0.037; effect size Cohen’s D=0.9). This spike-field coherence relationship did not exist for theta (z-scaled coherence for response neurons: 0.5(0.2-0.7) vs. non-response neurons 0.4(0.3-0.4); Wilcoxon rank sum *p*=0.29; effect size Cohen’s D=0.2). These data provide evidence that STN response-related activity could reflect frontal top-down novelty-induced orienting and control. Together, these data support the idea that novelty can trigger midfrontal low-frequency rhythms, which in turn engage subcortical STN circuits involved in response control.

**Figure 8.**
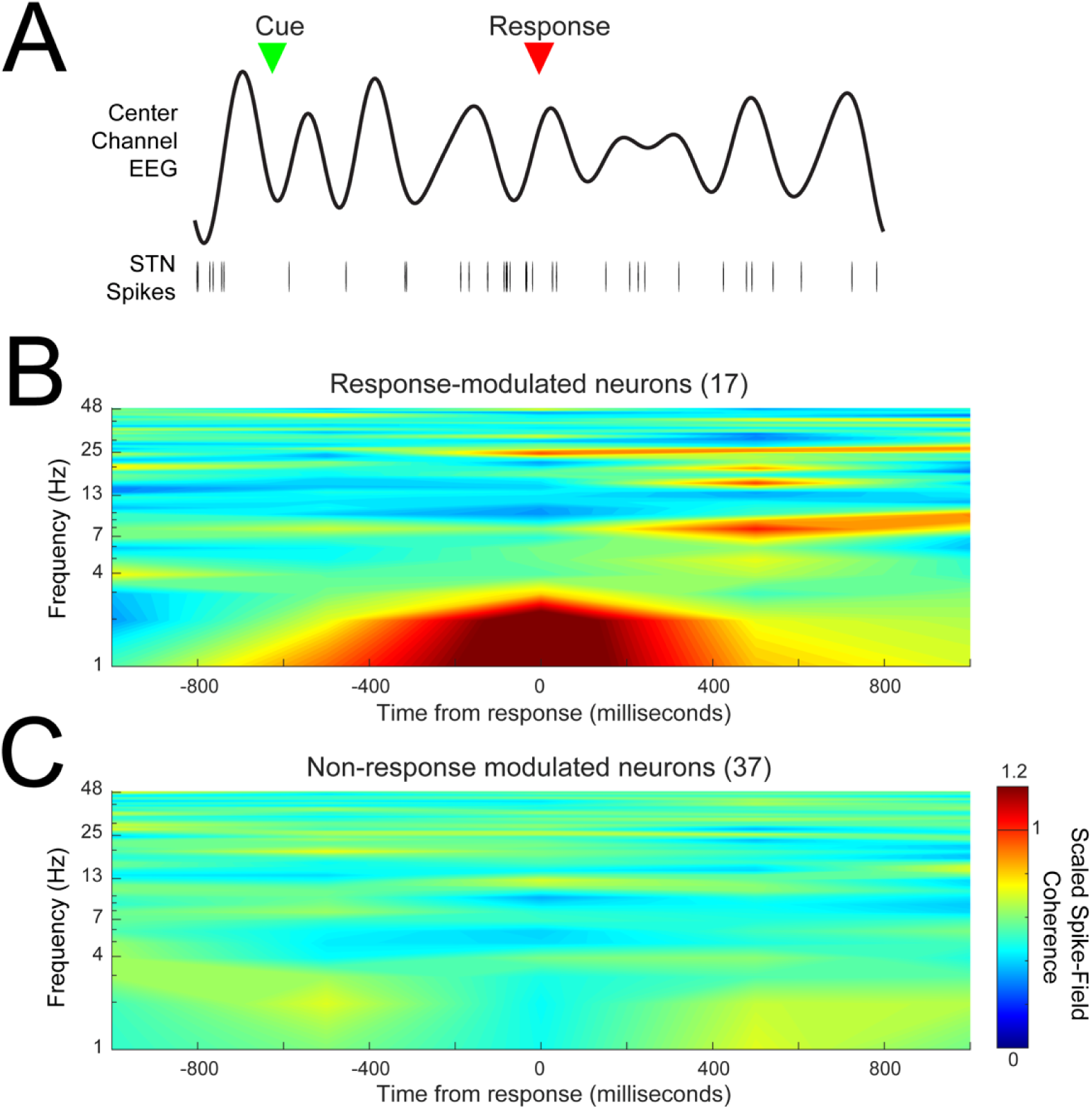
Response-related STN neurons had low frequency coherence with frontal EEG. A) An example time series from a single trial of center channel EEG and STN spiking activity; cue is denoted via a green triangle, and response is denoted via a red triangle. This neuron becomes coherent immediately with midfrontal low-frequency oscillations after the cue and prior to response. B) Across all 17 response-modulated STN neurons, there was significantly more 1-4Hz spike-field coherence with center-channel EEG electrodes than for C) 37 non-response modulated STN neurons. Spike-field coherence is scaled with 1 representing the 95% confidence-interval for coherence for comparison across neurons. Data from 54 STN neurons in 17 PD patient-volunteers.

## DISCUSSION

In this study, we combined scalp-based EEG and intraoperative recordings to examine novelty-related brain activity in frontal-subcortical circuits. We report three main results: 1) Both PD and control participants engaged low-frequency power in response to novel stimuli, with 1-4 Hz activity linked to novelty-related slowing whereas 4-7 Hz activity was specifically attenuated in PD participants, 2) STN neurons were modulated by novel responses, and 3) STN response-related neurons exhibited low-frequency coherence with midfrontal EEG activity. Our findings provide a mechanistic depiction of midfrontal-STN cognitive control systems in PD and neuronal responses to novel information. Taken together, our data support the idea that midfrontal low-frequency rhythms recruit subthalamic resources to slow, orient, and respond to new information.

Our work here is in line with past studies from our group and others showing that PD participants have attenuated midfrontal delta/theta rhythms between 1-7 Hz (Cavanagh et al., 2018; K.-H. Chen et al., 2016; Giller et al., 2020; Singh et al., 2018, 2021; Solís-Vivanco et al., 2018). This line of work includes data from the Simon reaction-time task (Singh et al., 2018), interval timing tasks (Singh et al., 2021), and working memory manipulations (Itthipuripat et al., 2013). Our past work has found that midfrontal delta 1-4 Hz frequencies are attenuated in PD (Kim et al., 2017; Parker et al., 2015) and correlated with cognitive function and behavior during an interval timing task (Singh et al., 2021). In this present study, we find that these rhythms are linked with novelty-related slowing and engaged with subcortical STN neurons, but not specifically attenuated in PD. By contrast, we found that novelty-triggered midfrontal 4-7 Hz oscillations were attenuated in PD participants, and not reliably linked with novely-related slowing, although we note that the relationship is in the same direction, and it is possible with more statistical power this relationship would be more consistent. Of note, there are multiple generators of mid-frontal low-frequency activity (Zuure et al., 2020), and it is possible that lower frequencies (1-4 Hz) are more consistently engaged by novelty-related slowing (Cavanagh et al., 2018; Lavallee et al., 2014; Parker et al., 2015; Wessel et al., 2016) while higher frequencies (4-7 Hz) may be more reliably impaired in PD (Cavanagh & Frank, 2014; Cohen & Donner, 2013; Singh et al., 2018; Töllner et al., 2017; Zuure et al., 2020).

We found that response-modulated STN neuronal activity was coherent with these lower frequencies (1-4 Hz). Our work is consistent with prior studies of STN neural activity; indeed, Bockova and colleagues (2011) found that the STN was modulated by distractor stimuli with a positive ERP peak around 200 ms, and Brittain and colleagues (2012) found that response inhibition is associated with STN activity (see also Alegre et al., 2013), which serves to suppress motor-related output from the basal ganglia. The low-frequency coherence we found is consistent with recent studies that revealed that oscillations in the STN during conflict are driven by midfrontal activity up to 7 Hz (Cavanagh et al., 2011, 2017; Zavala et al., 2014; Zeng et al., 2021). It is possible that multiple cortical 4-7 Hz theta features could couple with lower-frequency (∼2 Hz) STN oscillations engaging novelty. This could suggest that the cortex has more sophisticated information integration whereas the STN could more simply carry out reactive responses to novelty (e.g. stop & orient). Multiple studies have indirectly demonstrated connectivity between the frontal cortex and the STN in a pathway known as the hyperdirect pathway (Brunenberg et al., 2012; W. Chen et al., 2020; Haynes & Haber, 2013; Kelley et al., 2018), and midfrontal-STN activity may interact either directly via hyperdirect or via other key structures (such as the the thalamus) that are part of canonical basal ganglia circuits.

Our data help us understand how PD impacts novelty-related responses. Previous studies have shown that individuals with PD present with impaired habituation over repeated stimulus presentations, and that attenuated midfrontal theta activity is related to the rate of startle habituation (Cavanagh et al., 2018; K.H. Chen et al., 2016). Importantly, EEG habituation to novelty can effectively classify PD patients (Cavanagh et al., 2018). In our study, PD participants were not as successful as controls at quickly reorienting to the task at hand when presented with distracting novel stimuli, though we note that both controls and PD participants experienced an expected decrease in novelty-related slowing across the task. We found that PD participants also demonstrated reduced novelty-related frontal theta oscillations compared to controls (although theta power, unlike delta power, was not directly related to behavioral response speed). These findings suggest that structural and functional changes related to PD affect the circuitry that evaluates and responds to novel stimuli.

Our work has several limitations. First, intraoperative recordings present many challenges, including a lack of control over the experimental environment. In particular, intraoperative conditions are quite different from scalp EEG sessions; in addition, we used adhesive electrodes placed somewhat anterior to the peak of midfrontal ∼4-Hz activity our whole scalp EEG experiment captured (i.e. **Figure 2E**). These factors may have affected our novelty-related findings in **Figure 4C**. Because STN single-unit recordings occur in PD patients, it is unclear how our STN findings generalize to non-PD patients or patients with other brain diseases that disrupt frontostriatal circuits.

Overall, we found evidence for a ∼4-Hz stimulus-response arc between the frontal cortex and STN during novelty. Our findings align with recent work demonstrating clinical relevance of midfrontal ∼4-Hz activity in PD (Singh et al., 2021; Solís-Vivanco et al., 2018). Further, small studies have revealed that STN low-frequency (∼4–5 Hz) stimulation can beneficially impact conflict (Scangos et al., 2017) and interval timing (Kelley et al., 2018), as well as oscillatory activity in the prefrontal cortex (Bentley et al., 2020). These findings in humans are further supported by evidence from animal models of PD showing that highly-specific low-frequency stimulation can improve interval timing (Kim et al., 2017; Kim & Narayanan, 2019). Restoring behaviorally-relevant ∼4-Hz oscillations may contribute directly to improved cognitive performance. Future research should systematically evaluate this mechanism in humans.

Overall, the current studies show that novelty-related distraction is more evident in individuals with PD compared to controls. We find that that low-frequency delta power is related to novelty-related slowing and individuals with PD had decreased novelty-responsive midfrontal 4-7 Hz rhythms compared to controls. Furthermore, we find that neurons in the STN are modulated around responses to novelty, and that neurons in the STN and the midfrontal cortex have low-frequency coherence around responses to novelty. Our data illuminate how novelty modulates ∼4-Hz rhythms in frontal-STN circuits, which may provide insight into neuronal responses to novelty and PD-related changes in cognitive control. This work may be significant in the development of novel biomarkers or treatments for PD and other brain diseases degrading basal ganglia circuits.

## Data Availability

Data is available at narayanan.lab.uiowa.edu

https://narayanan.lab.uiowa.edu/

## Contributions

RC, NN, JFC, and JG conceived and designed experiments; RC and AE recorded EEG data; RC and JG performed intraoperative recordings; AS, RC, JFC, and NN wrote analysis code; RC, JFC, NN, and JG wrote the paper; and JB independently checked the data.

## Acknowledgements

We would like to acknowledge Haiming Chen, Youngcho Kim, Derrick Okine, Darcy Diesberg, and Tobin Dykstra for technical contributions and helpful discussions.

